# The NUTRIENT Trial (NUTRitional Intervention among myEloproliferative Neoplasms): Feasibility Phase

**DOI:** 10.1101/2023.05.09.23289740

**Authors:** Laura F. Mendez Luque, Julio Avelar-Barragan, Hellen Nguyen, Jenny Nguyen, Eli M. Soyfer, Jiarui Liu, Jane H. Chen, Nitya Mehrotra, Heidi E. Kosiorek, Amylou Dueck, Alexander Himstead, Elena Heide, Melinda Lem, Kenza El Alaoui, Eduard Mas Marin, Robyn M. Scherber, Ruben A. Mesa, Katrine L. Whiteson, Andrew Odegaard, Angela G. Fleischman

## Abstract

**Purpose:** Chronic inflammation is integral to Myeloproliferative Neoplasm (MPN) pathogenesis. JAK inhibitors reduce cytokine levels, but not without significant side effects. Nutrition is a low-risk approach to reduce inflammation and ameliorate symptoms in MPN. We performed a randomized, parallel-arm study to determine the feasibility of an education-focused Mediterranean diet intervention among MPN patients.

**Experimental Design:** We randomly assigned participants to either a Mediterranean diet or standard US Dietary Guidelines for Americans (USDA). Groups received equal but separate education with registered dietician counseling and written dietary resources. Patients were prospectively followed for feasibility, adherence, and symptom burden assessments. Biological samples were collected at four time points during the 15-week study to explore changes in inflammatory biomarkers and gut microbiome.

**Results:** The Mediterranean diet was as easy to follow for MPN patients as the standard USDA diet. Over 80% of the patients in the Mediterranean diet group achieved a Mediterranean Diet Adherence Score of ≥8 throughout the entire active intervention period, whereas less than 50% of the USDA group achieved a score of ≥8 at any time point. Improvement in symptom burden was observed in both diet groups. No significant changes were observed in inflammatory cytokines. The diversity and composition of the gut microbiome remained stable throughout the duration of the intervention.

**Conclusions:** With dietician counseling and written education MPN patients can adhere to a Mediterranean eating pattern. Diet interventions may be further developed as a component of MPN care, and potentially even be incorporated into the management of other chronic clonal hematologic conditions.

**STATEMENT OF TRANSLATIONAL RELEVANCE:** Chronic clonal hematologic disorders, such as myeloproliferative neoplasm (MPN), lie at the intersection between malignancy and chronic inflammatory disease. Chronic inflammation is responsible for many of the clinical consequences of MPN. Diet is a central tenant of management of chronic conditions characterized by subclinical inflammation, such as cardiovascular disease, but has not entered the treatment algorithm for clonal hematologic disorders. Here, we establish that a Mediterranean diet intervention is feasible in the MPN patient population and can improve symptom burden. These findings warrant large dietary interventions in patients with clonal hematologic disorders to test the utility of diet in improvement of clinical outcomes.

## INTRODUCTION

Myeloproliferative neoplasms (MPN), including polycythemia vera (PV), essential thrombocythemia (ET), and primary myelofibrosis (PMF), are hematologic malignancies characterized by the clonal outgrowth of hematopoietic cells with a somatically acquired mutation most commonly in *JAK2* (*JAK2*^*V617F*^)(1-5). The clinical consequences of MPN include thrombosis, transformation to acute leukemia, abnormal blood counts, and a significant symptom burden. MPN is a highly inflammatory disease, with increased plasma cytokines as a hallmark feature of the disease.

Chronic inflammation is pervasive in MPN, contributing to symptomatology(6), blood count abnormalities(7), and disease progression(8,9). JAK inhibitors reduce inflammation(10,11), resulting in amelioration of symptoms and improvement in quality of life(12,13). However, JAK inhibitors are not without risks including immunosuppression, weight gain(14) and skin cancers(15), are extremely costly, and are not indicated for all MPN patients. Recently developed National Comprehensive Cancer Network (NCCN) guidelines for MPN address the importance of symptom burden, and recommend intervention to reduce symptom burden regardless of prognosis scoring category(16). However, many MPN patients do not meet criteria for a cytoreductive agent. Therefore, many MPN patients are maintained without intervention that adequately address symptoms nor impact disease progression. Lifestyle modifications to reduce inflammation, such as diet, could have significant short term as well as long term positive impacts on the disease. In the short term, adopting a healthful diet rich in anti-inflammatory foods may serve to improve symptom burden among MPN patients. In the long-term minimizing inflammation through diet may potentially delay or prevent disease progression.

The Mediterranean diet, characterized by increased consumption of extra virgin olive oil (EVOO), nuts, legumes, vegetables, fruits, fish, and whole grain products, has proven to be beneficial in diseases where chronic subclinical inflammation plays a key role(17). For example, the PREDIMED (Prevención con Dieta Mediterránea) study demonstrated that a Mediterranean diet supplemented with EVOO or nuts reduced the incidence of major cardiovascular events(18). The Mediterranean diet’s anti-inflammatory properties are attributed to its richness in phenolic compounds and nutrient density(19).

Data is emerging that dietary and microbiome factors may be beneficial in clonal hematologic disorders(20,21). In a cohort of multiple myeloma patients on lenalidomide maintenance consumption of dietary flavonoids correlated with stool butyrate concentration, and higher stool butyrate concentration was associated with sustained Minimal Residual Disease (MRD) negativity(21). An ongoing dietary intervention study (NUTRIVENTION) of a whole food plant-based diet in patients with monoclonal gammopathy and smoldering multiple myeloma will evaluate the impact of diet in these precursor conditions(22).

Nutritional control of inflammation represents a unique low risk therapeutic approach to alleviate the symptom burden of MPN patients and to also possibly blunt disease progression. Here, we investigated the feasibility of employing an education-focused Mediterranean diet intervention in an MPN patient cohort. Our main goal was to establish that MPN patients are willing and able to initiate dietary education as a potential symptom burden management. Further, we also collected preliminary efficacy and mechanistic data on symptom burden, inflammatory cytokines, and the gut microbiome.

## METHODS

### Study design and participants

The NUTRIENT study was a single center interventional pilot study of an educational dietary intervention among MPN patients performed at University of California, Irvine from October 2018 through December 2019. The protocol was approved by the IRB of University of California Irvine and registered on clinicaltrials.gov (NCT04744974). The study was conducted in accordance with the U.S. Common Rule.

### Endpoints

We had a combined primary endpoint of both feasibility of and adherence to a Mediterranean diet assessed via online surveys. Feasibility was assessed via a single-item question on each of the online surveys administered while participants were actively receiving intervention of “how easy do you feel this diet is to follow?” on a 0 to 10 numerical score (0 very easy to 10 very difficult) with a score of <5/10 being regarded as reasonably easy to follow. Adherence was assessed using the 14-point Mediterranean Diet Adherence Screener (MEDAS) with slight modifications in how the questions were worded. We defined good adherence to a Mediterranean style eating pattern as a score of ≥ 8 on the MEDAS, which is the top tertial and has been used as a benchmark for good adherence in other US based studies(23). As a second mode of dietary assessment participants completed online 24-hour diet recalls using the National Cancer Institute Automated Self-Administered 24-hour (ASA24®) dietary assessment tool at unannounced days during weeks 1, 2, 3, 6, 9, 12, and 15 which was used to calculate the United States Dietary Guidelines for Americans (USDA) Healthy Eating Index 2015 score (HEI-2015)(24). Exploratory endpoints included plasma concentration of inflammatory cytokines, reduction in symptom burden, changes in hematologic parameters, lipids, and change in the gut microbiome.

### Study Schedule

The total duration of the study was 15 weeks (Figure 1). During weeks 1-2 participants were followed without intervention, during which we obtained two baseline measures of dietary intake and symptom burden (one at enrollment and a second unannounced one during the two week lead in time) and one biological sample (blood and stool). Participants received a total of 10 weeks active dietary intervention (week 3-12) with an in-person meeting with a dietician and distribution of dietary resources, during which four surveys, four dietary recalls, and two biological samples were collected. During weeks 13-15 participants no longer received weekly educational materials, at week 15 participants completed one survey, one 24-hour dietary recall, and contributed one set of biological samples (blood, stool).

**Figure 1.**
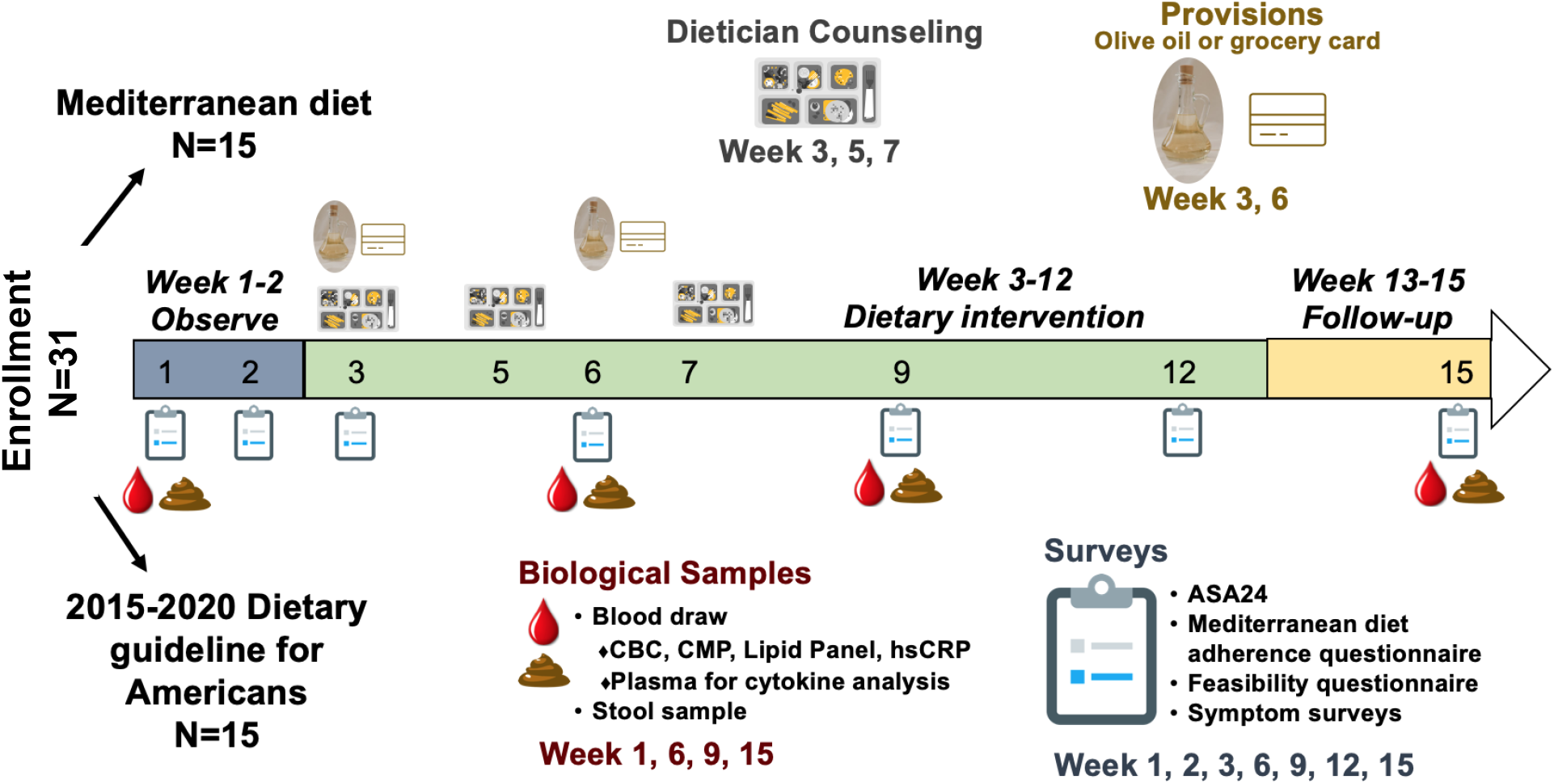
NUTRIENT study design

### Intervention

All participants met once at the start of the intervention period (week 3) with a registered dietician for one-on-one counseling to educate the participant on the central components of the Mediterranean diet or the US Dietary guidelines, and to tailor the diet to meet each participant’s medical needs and/or cultural preferences. Participants had two follow up dietary counseling visits during week 5 and 7. Participants were emailed 10 weekly installments of educational materials on their respective diet in a pdf format during week 3-12. All participants in the Mediterranean diet arm were given 750 milliliters of extra virgin olive oil (EVOO) at week 3 and 6, and all participants following the standard US Guidelines diet were given a $10 grocery gift card at week 3 and 6. During this time period participants completed four online surveys (week 3, 6, 9, 12) and donated two sets of blood, stool, and urine samples (weeks 6, 9).

### Data Collection

#### Laboratory Studies

Four biological sample data points were collected during the 15-week study which included collection of blood, stool, and urine. Peripheral blood was collected at week 1, 6, 9, and 15, complete blood count (CBC), comprehensive metabolic panel, lipid panel, and high sensitivity C reactive protein (hs-CRP) were performed by the clinical laboratory of UCI Health. Blood was centrifuged within 2 hours of draw to obtain plasma and stored at ^-^80°C for cytokine measurements.

#### Cytokine Analysis

Frozen plasma was performed by Quanterix in Billerica, MA for analysis. A Human CorPlex 10 Cytokine Array kit #85-0329 (IL-12p70, IL-1B, IL-4, IL-5, IFN?, IL-6, IL-8, IL-22, TNFα, and IL-10) was used according to manufacturer’s protocol and analyzed using a Quanterix SPX imager system on-site at Quanterix Headquarters in Billerica, MA. Cytokines were transformed to a log base 2 scale for analysis purposes. Mixed models (with a random intercept for each participant) were used to explore changes over time where group and time were fixed effects and an interaction term was included.

#### Collection of fecal samples

Study participants provided a fecal sample at 4 different timepoints (weeks 1, 6, 9, and 15) stored in Zymo DNA/RNA shield preservation buffer (Cat. #R1101). These were returned in person or by mail. Samples were then stored at ^-^80°C until analysis.

#### Extraction of DNA from fecal samples

DNA was extracted from feces by thawing the samples on ice and homogenizing them. Afterwards, 1 mL of the fecal slurry was extracted using the ZymoBiomics DNA Miniprep Kit (Cat. #D4300) in accordance with the manufacturer’s suggested protocol. Bead lysis during the extraction was performed at 6.5 m/s for 5 minutes total using a MPBio FastPrep-24 instrument.

#### Library preparation and sequencing

Libraries for shotgun metagenomic sequencing of extracted fecal DNA were prepared using the Illumina DNA prep kit (Cat. # 20018705), using an adapted low-volume protocol(25). DNA quantification of the final library pool was performed using the Quanti-iT PicoGreen dsDNA kit (Cat. #P7589). Synthetic microbial DNA standards were included as positive sequencing controls (ZymoBIOMICS Microbial Community DNA Standard, Cat. #D6305), and PCR grade water was used as a negative sequencing control. Sequencing was performed by Novogene Corporation Inc. (Sacramento, CA) using Illumina’s Hiseq 4000. An average of 2,819,107 +/- 670,543 paired-end reads per sample, 150 bases in length, were obtained. Data is available on the Sequence Read Archive under the BioProject ID, PRJNA918651.

#### Microbiome analysis

First, raw sequencing data was quality filtered, and host-derived reads were removed. Taxonomic assignment of sequences was performed using MetaPhlAn3 and its default parameters(26). A table of species relative abundances per sample was produced and subsequently analyzed in R v4.2.1. The Shannon diversity index, Bray-Curtis dissimilarity matrix, and principal coordinate ordination were performed using the Vegan v2.5-6 package in R(27). Significance testing of microbiome diversity and composition metrics was done using linear-mixed effect models in the nlme v3.1-148 package(28). Further details regarding the analysis of microbiomes can be found in our companion manuscript(29). The code used for this analysis is available at https://github.com/Javelarb/MPN_diet_intervention.

## RESULTS

### Purpose of Study

The primary objective of the NUTRIENT study was to assess whether MPN patients can adopt a Mediterranean eating pattern with dietician counseling and educational materials. Thirty-one MPN patients were randomly assigned to receive either dietician counseling and written educational materials on the Mediterranean Diet (MED) or the 2015-2020 United States Dietary Guidelines for Americans (USDA), 28 participants completed the study. Participants were told they would be randomized to one of two diets that are conventionally regarded to be healthful but were not informed of the specific diets being studied nor which group they were randomized to. The US Dietary Guidelines for Americans was chosen as an intervention that would provide the participants with equal counseling attention but did not encourage a Mediterranean diet eating pattern.

### Patient Recruitment and Demographics

We recruited individuals who were 18 years of age or older and who had been previously diagnosed with a Philadelphia chromosome negative MPN including essential thrombocythemia (ET), polycythemia vera (PV), or myelofibrosis (MF, includes primary myelofibrosis as well as post-ET or post-PV myelofibrosis). Any type of MPN directed therapy was allowed. A complete list of inclusion and exclusion criteria are provided in Supplemental Table 1. We screened 47 potential participants. Thirty-one participants were randomized, two withdrew due to family illness and one was lost to follow-up. Demographics of the 28 patients who completed the study are shown in Table 1.

**Table 1.** Demographics of study cohort

|  | **USDA (n=13)** | **Mediterranean Diet (n=15)** |
| --- | --- | --- |
| **Female n (%)** | 10 (76%) | 10 (67%) |
| **Age mean (range)** | 58 (21-77) | 57 (25-71) |
| **Disease n (%)** |  |  |
| PV | 6 (46%) | 8 (53%) |
| ET | 3 (23%) | 3 (20%) |
| MF | 4 (31%) | 4 (27%) |
| **Mutation n (%)** |  |  |
| JAK2 | 12 (92%) | 13 (87%) |
| CALR | 0 | 2 (13%) |
| MPL | 1 (8%) | 0 |
| **Treatment n (%)** |  |  |
| Hydroxyurea | 4 (31%) | 5 (34%) |
| Ruxolitinib | 1 (8%) | 2 (13%) |
| Interferon | 2 (15%) | 3 (20%) |
| Other | 6 (46%) | 5 (33%) |
| **MPN-SAF median (interquartile range, IQR)** | 15.43 (17.15) | 10.29 (12.15) |
| **Mediterranean Diet Adherence Score prior to starting intervention median (IQR)** | 6.50 (1.71) | 8.46 (1.57) |

### Adoption of a Mediterranean Eating Pattern

Adherence to a Mediterranean style eating pattern was assessed at weeks 1, 2, 3, 6, 9, 12, and 15 using a 14-point Mediterranean Diet Adherence Score (MEDAS)(30). Because of inconsistent timing of the week 3 surveys with relation to the initial dietician visit this timepoint was removed from the analysis. A MEDAS score of ≥8 was defined as having adequate adherence to a Mediterranean style eating pattern(23). Our pre-defined goal was to have at least 80% of the participants in the Mediterranean group maintaining a MEDAS score of ≥8 during the active intervention period. At all timepoints during the active intervention (weeks 6, 9, 12) ≥80% of the MED group maintained a MEDAS score of ≥8, at no time point did at least 80% of the USDA group achieve a MEDAS score of ≥8 (Figure 2A).

**Figure 2.**
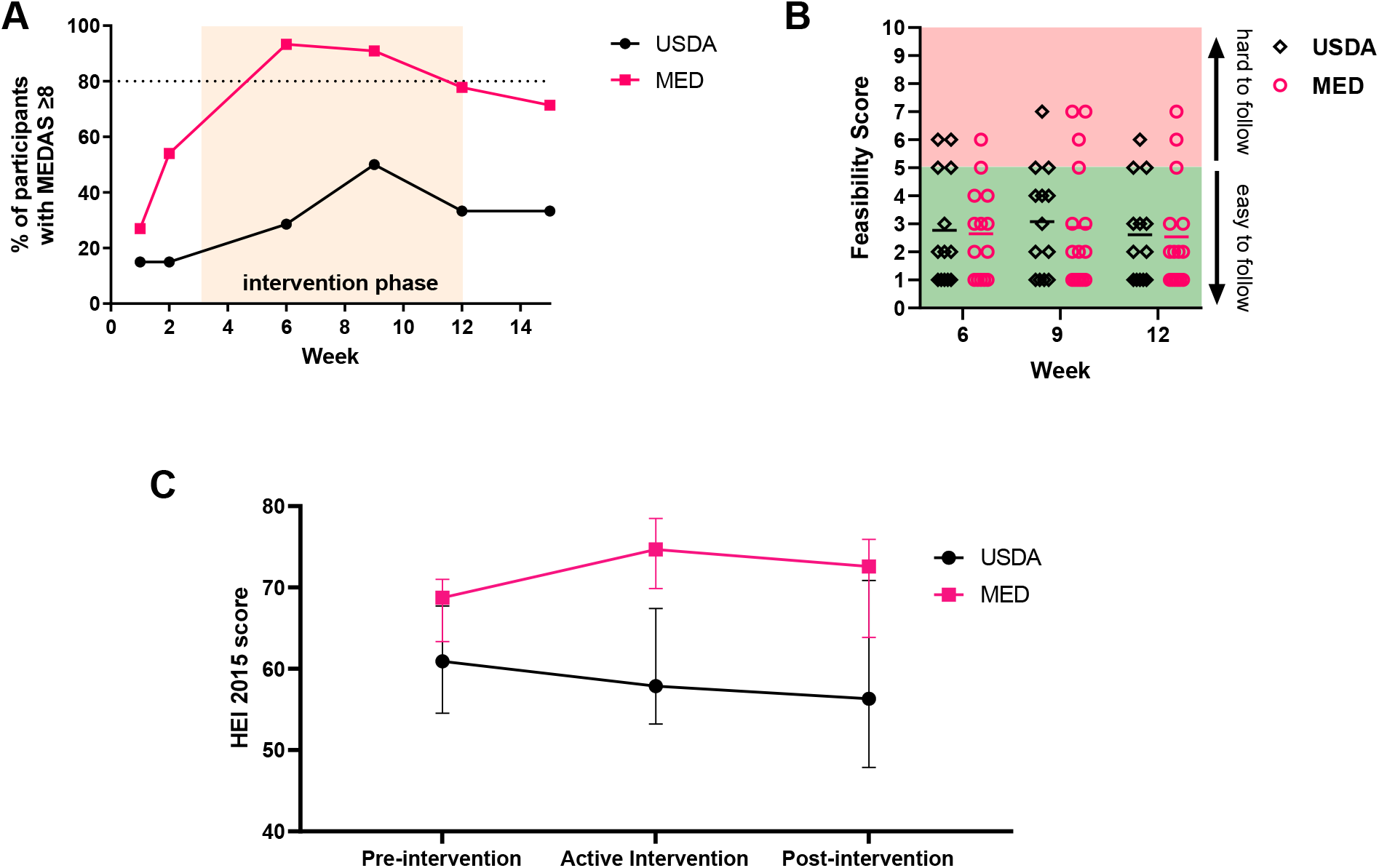
MPN patients can adopt a Mediterranean eating pattern with dietician counseling and education. (A) Percentage of participant with MEDAS scores ≥8 at each time point with orange shaded area depicting the active intervention period (B) Participant responses to feasibility question during active intervention period (C) HEI-2015 was calculated from each 24 hour diet recall, and scores for each participant were averaged for the pre-intervention (weeks 1-2), active intervention (weeks 3-12), and post-intervention (weeks 13-15) period. Data shown represents median with interquartile range.

We assessed feasibility with a single item question in each survey asking “how easy do you feel this diet is to follow”, we pre-defined a feasibility benchmark as at least 3 of the 4 assessments achieving a score of <5/10 on a 0-10 numerical score. Seventy percent of the patients on the USDA diet achieved the pre-defined feasibility benchmark, and 79% of patients in the Mediterranean arm achieved this feasibility benchmark (Figure 2B). This demonstrates that a Mediterranean diet is at least as easy to follow for MPN patients as the standard US Dietary Guidelines for Adults.

### Diet Quality of Participants

We used the 24 hour diet recall data (ASA24®) to calculate the Healthy Eating Index (HEI-2015)(24) as a measure of general diet quality. The median HEI-2015 score for the MED group rose from 69 pre-intervention to 75 during the active intervention and maintained at 73 post-intervention. The median HEI-2015 scores for the USDA group was 61 pre-intervention, 58 during the active intervention, and 56 post-intervention (Figure 2C).

### Impact of diet on symptom burden

Symptom burden was assessed using the MPN-SAF TSS (also known as MPN-10), which grades the 10 most clinically relevant symptoms of MPN patients(31). We supplemented this questionnaire with extended gastrointestinal focused symptoms. Approximately half of the Mediterranean diet cohort enjoyed a > 50% reduction in MPN-TSS at weeks 9-15 compared to baseline, whereas the maximum percentage of participants in the USDA arm achieving > 50% reduction in their MPN-TSS was 31% at 15 weeks (Figure 3A). We also calculated the mean change in each specific symptom queried on the surveys to visualize the impact of the diets on specific symptoms (Figure 3B).

**Figure 3.**
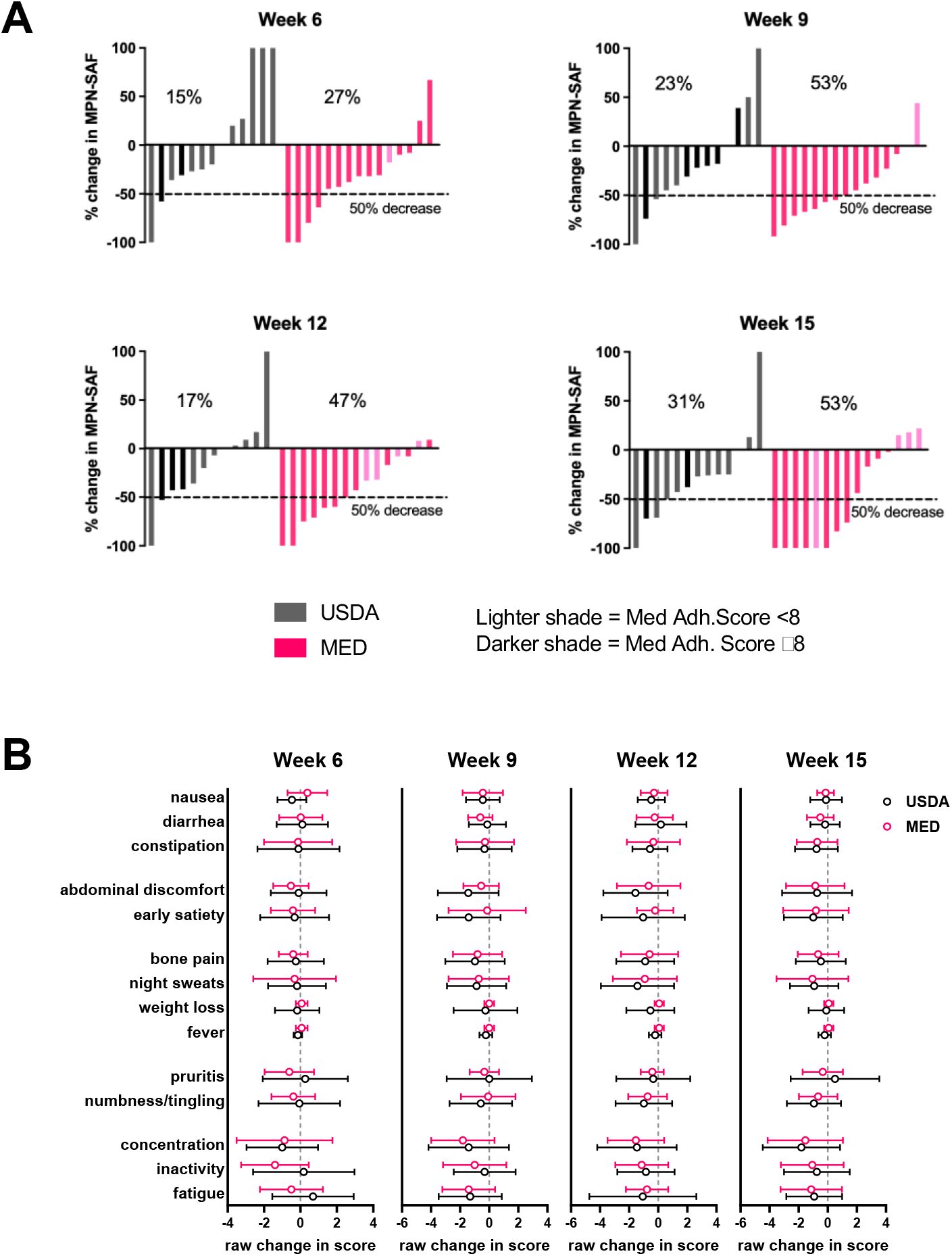
Changes in symptom burden during study. (A) waterfall plots of percentage change in MPN-SAF (MPN-TSS) at each week compared to baseline (baseline defined as average MPN-TSS of weeks 1 and 2) (B) Raw change in specific symptoms at each week compared to baseline (mean±SD).

### Impact of Diet on Laboratory Parameters

Laboratory data, including complete blood counts with differential (CBC w/diff) (Supplemental Figure 1), comprehensive metabolic panel (CMP) (Supplemental Table 2), and lipid profiles (Supplemental Table 3) were collected at week 1 (pre-intervention), weeks 6 and 9 (during active intervention), and week 15 (post-intervention). Blood counts, kidney function, and liver function remained stable during the intervention, demonstrating that a diet intervention is safe in the MPN patient population.

### Impact of diet on inflammatory biomarkers

We explored the impact of the diet intervention on biological measures of inflammation, including high sensitivity C-reactive protein (hsCRP) (Supplemental Figure 2) and plasma cytokines. Out of the ten cytokines measured, five cytokines (TNFα, IL-6, IL-8, IL-10, IL-22) yielded levels within the detectable range (Figure 4). In these five cytokines there were no significant differences in changes over time by group.

**Figure 4.**
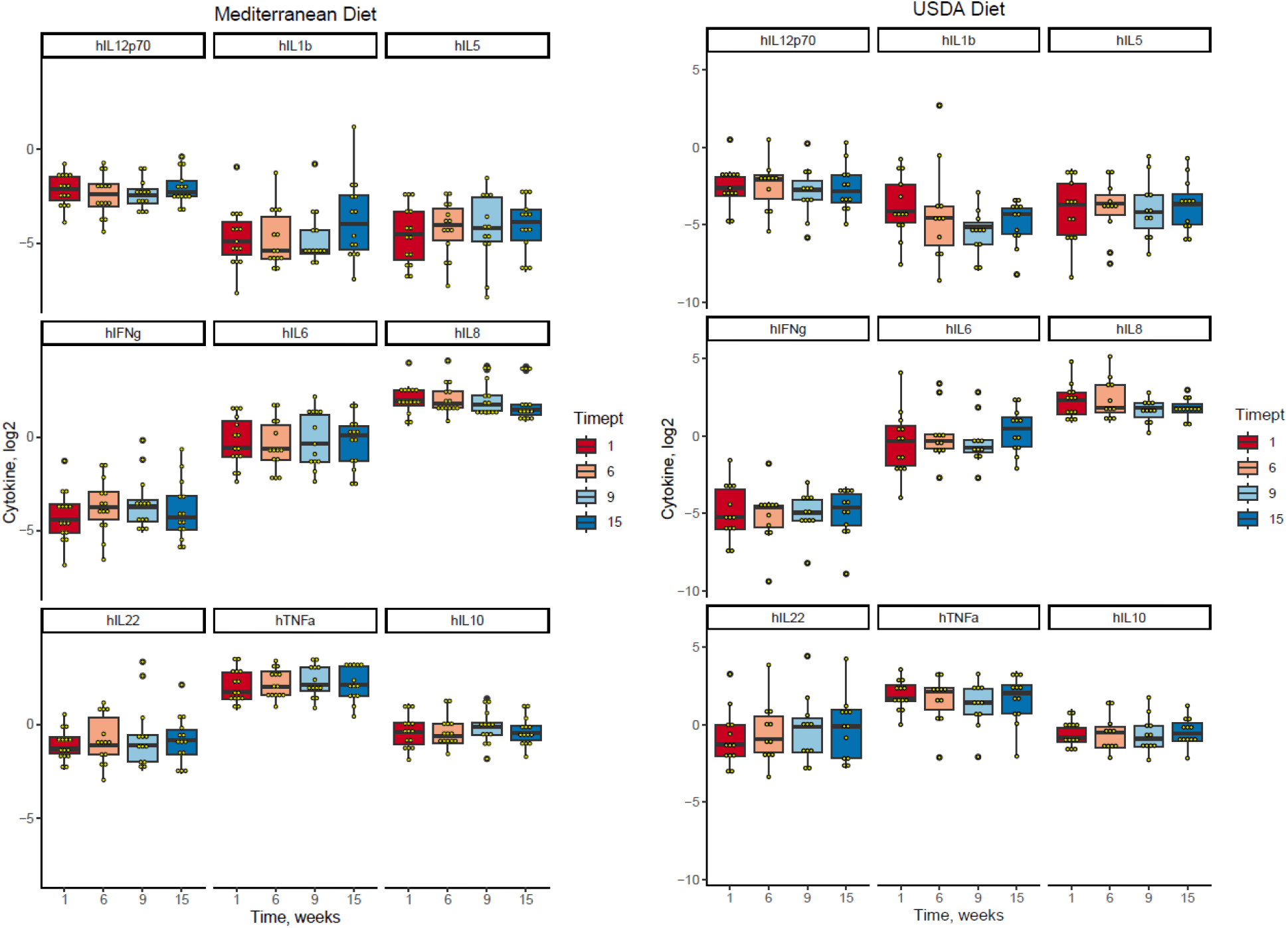
Changes in plasma cytokine concentrations throughout the study. Plasma cytokines were measured at weeks 1, 6, 9, and 15.

### Impact of diet on gut microbiome

We used shotgun metagenomic sequencing to evaluate the gut microbiome at weeks 1 (pre-intervention), weeks 6 and 9 (during active intervention), and week 15 (post-intervention). We found that microbiome diversity and composition was stable throughout the study duration in both cohorts, with no differences in the Shannon diversity index (p = 0.57) or first principal coordinate of microbiome composition (p = 0.25) due to diet (Figure 5). A more detailed analysis of the microbiome of this cohort and correlations with cytokines are described in a separate manuscript (Avelar-Barragan et al, in preparation, *https://doi.org/10.1101/2023.01.25.525620*).

**Figure 5:**
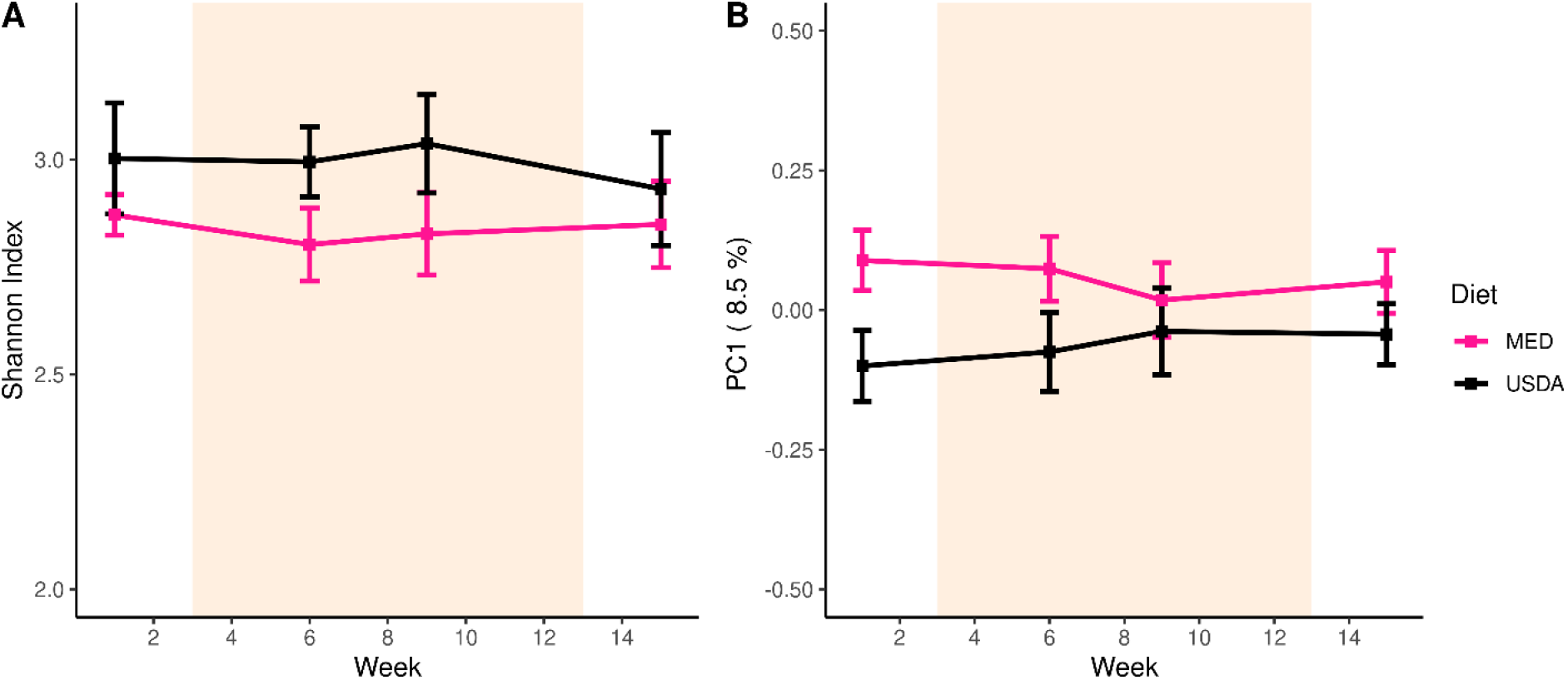
Fecal Microbiome Composition. (A) A line plot displaying the microbial diversity, as measured by the Shannon Index, of individuals over time. (B) A line plot displaying the microbial composition of individuals over time. The y-axis is the first principal coordinate produced by Bray-Curtis dissimilarity ordination of the microbiome. In both (A) and (B), the orange shaded area depicts the active intervention period and the standard error is represented by error bars.

### MPN driver mutation allele burden

We quantified the *JAK2*^*V617F*^ allele burden of JAK2-positive patients from whole blood during the trial using digital PCR. We observed minimal changes of the *JAK2*^*V617F*^ allele burden in both diet groups (Supplemental Figure 3) over the short duration of the study.

## DISCUSSION

The central purpose of the NUTRIENT trial was to establish the feasibility of diet as a therapeutic approach in MPN. Diet may be a low-risk, low-cost approach to reduce inflammatory cytokines thus alleviating symptoms and potentially preventing disease progression. In addition, empowering MPN patients to take an active role in their treatment through diet may instill a greater sense of well-being and improve quality of life.

The Mediterranean diet was chosen because it is generally accepted to be a healthful diet, is rich in anti-inflammatory and antioxidant compounds, and has been found to reduce inflammatory biomarkers(19). MPN patients reported that a Mediterranean diet program was just as easy to follow as a program based on the US Guidelines for Americans (Figure 2). Using the MEDAS as the primary measure to quantify adherence to a Mediterranean style eating pattern we found that ≥80% of participants in the Mediterranean diet group were able to maintain good adherence to a Mediterranean diet eating pattern throughout the active intervention period, compared to less than 50% of the participants in the USDA group. This demonstrates that MPN patients can adopt a Mediterranean diet eating pattern with dietician counseling and written curriculum. Development of an MPN specific Mediterranean diet curriculum tailored to address specific issues in this population such as early satiety, fatigue, and reduction of iron in Polycythemia Vera (PV) could improve feasibility and enhance adherence further.

Targeting symptoms is an important goal in MPN, as symptoms negatively impact quality of life, increase use of medical care, and result in loss of productivity. Although not statistically powered to detect a change, we explored the impact of a diet intervention on symptom burden. A general improvement in diet quality would be expected to lead to an improvement in well-being which would translate into a reduction in symptom score. Participants in both groups enjoyed a reduction in symptom burden. In the USDA group 15, 23, 17 and 31% had a >50% reduction in their MPN-TSS score at 6, 9, 12, and 15 weeks, respectively. In the MED group 27, 53, 47, and 53% had a > 50% reduction in their MPN-TSS at 6, 9, 12, and 15 weeks. This data suggests that a general improvement in diet quality can impact symptoms, but that the components of a Mediterranean diet may augment symptom improvement.

The length of the diet intervention and intensity of sessions may be an important factor in creating a change in eating habits of participants. Our active intervention period consisted of 10 weeks, with an additional 2 week lead in time and 3 weeks post-intervention follow up. For dynamic endpoints such as symptom burden it appears that a 10-week intervention is sufficient to detect change. However, with a longer intervention period one could examine whether there is sustained improvement in symptoms. A longer intervention period may also identify a subset of people with delayed symptom improvement, potentially revealing that symptoms stem from different root causes, some of which are quickly changed and others of which take some time to change.

The lack of identifying a reduction in inflammatory cytokines was not surprising given our small sample size. Future larger studies are required to assess the impact on inflammatory cytokines, in addition potentially longer time periods are needed to observe decreases in inflammatory cytokines. Or, perhaps alternative approaches to capture and quantify inflammation are required.

This trial afforded us the opportunity to explore potential changes in the gut microbiome over time with a diet intervention. Although the microbiome remained stable throughout the study in both cohorts, a Mediterranean diet has been shown to beneficially alter the gut microbiome over time(32). Additionally, the polyphenolic compounds found in EVOO select for microbes associated with reduced inflammation(33,34). This suggests that adherence to a MED diet may indeed be impactful on the gut microbiome of MPN patients and is worthwhile to evaluate in a larger diet intervention cohort.

A Mediterranean diet intervention is feasible in the MPN patient population. This population is receptive to using diet as a therapeutic approach and can alter their diet toward a Mediterranean diet eating pattern. The benefits of diet may be seen most when the intervention is started early, for example in low-risk Polycythemia Vera or Essential Thrombocythemia patients who are commonly not given MPN specific therapy. Diet may also be a useful intervention in precursor conditions such as clonal hematopoiesis of indeterminate potential (CHIP), acting not only to ameliorate the negative health consequences of CHIP but to also prevent progression to hematologic malignancy.

## Data Availability

All data produced in the present study are available upon reasonable request to the authors

https://github.com/Javelarb/MPN_diet_intervention

## REFERENCES

1. Baxter EJ, Scott LM, Campbell PJ, East C, Fourouclas N, Swanton S, et al. Acquired mutation of the tyrosine kinase JAK2 in human myeloproliferative disorders. Lancet (London, England) 2005;365(9464):1054–61.

2. James C, Ugo V, Le Couédic J-P, Staerk J, Delhommeau F, Lacout C, et al. A unique clonal JAK2 mutation leading to constitutive signalling causes polycythaemia vera. Nature 2005;434(7037):1144–8.

3. Kralovics R, Passamonti F, Buser AS, Teo S-S, Tiedt R, Passweg JR, et al. A gain-of-function mutation of JAK2 in myeloproliferative disorders. The New England journal of medicine 2005;352(17):1779–90.

4. Levine RL, Wadleigh M, Cools J, Ebert BL, Wernig G, Huntly BJ, et al. Activating mutation in the tyrosine kinase JAK2 in polycythemia vera, essential thrombocythemia, and myeloid metaplasia with myelofibrosis. Cancer cell 2005;7(4):387–97.

5. Zhao R, Xing S, Li Z, Fu X, Li Q, Krantz SB, et al. Identification of an acquired JAK2 mutation in polycythemia vera. The Journal of biological chemistry 2005;280(24):22788–92.

6. Geyer HL, Dueck AC, Scherber RM, Mesa RA. Impact of Inflammation on Myeloproliferative Neoplasm Symptom Development. Mediators Inflamm 2015;2015:284706 doi 10.1155/2015/284706.

7. Fleischman AG, Aichberger KJ, Luty SB, Bumm TG, Petersen CL, Doratotaj S, et al. TNFalpha facilitates clonal expansion of JAK2V617F positive cells in myeloproliferative neoplasms. Blood 2011;118(24):6392–8.

8. Craver BM, El Alaoui K, Scherber RM, Fleischman AG. The Critical Role of Inflammation in the Pathogenesis and Progression of Myeloid Malignancies. Cancers 2018;10(4) doi 10.3390/cancers10040104.

9. Fleischman AG. Inflammation as a Driver of Clonal Evolution in Myeloproliferative Neoplasm. Mediators of inflammation 2015;2015:606819 doi 10.1155/2015/606819.

10. Verstovsek S, Mesa RA, Gotlib J, Levy RS, Gupta V, DiPersio JF, et al. A double-blind, placebo-controlled trial of ruxolitinib for myelofibrosis. The New England journal of medicine 2012;366(9):799–807.

11. Harrison CN, Kiladjian J-J, Gisslinger H, Passamonti F, Sirulnik LA, Wang L, et al. Association of cytokine levels and reductions in spleen size in COMFORT-II, a phase III study comparing ruxolitinib to best available therapy (BAT). Journal of Clinical Oncology 2012;30(15_suppl):6625- doi 10.1200/jco.2012.30.15_suppl.6625.

12. Harrison CN, Mesa RA, Kiladjian J-J, Al-Ali H-K, Gisslinger H, Knoops L, et al. Health-related quality of life and symptoms in patients with myelofibrosis treated with ruxolitinib versus best available therapy. British journal of haematology 2013;162(2):229–39 doi 10.1111/bjh.12375.

13. Mesa RA, Gotlib J, Gupta V, Catalano JV, Deininger MW, Shields AL, et al. Effect of ruxolitinib therapy on myelofibrosis-related symptoms and other patient-reported outcomes in COMFORT-I: a randomized, double-blind, placebo-controlled trial. Journal of Clinical Oncology 2013;31(10):1285–92 doi 10.1200/JCO.2012.44.4489.

14. Molle N, Krichevsky S, Kermani P, Silver RT, Ritchie E, Scandura JM. Ruxolitinib can cause weight gain by blocking leptin signaling in the brain via JAK2/STAT3. Blood 2020;135(13):1062–6 doi 10.1182/blood.2019003050.

15. Blechman AB, Cabell CE, Weinberger CH, Duckworth A, Leitenberger JJ, Zwald FO, et al. Aggressive Skin Cancers Occurring in Patients Treated With the Janus Kinase Inhibitor Ruxolitinib. Journal of drugs in dermatology : JDD 2017;16(5):508–11.

16. NCCN Clinical Practice Guidelines in Oncology (NCCN Guidelines®) Myeloproliferative Neoplasms Volume Version 2.20222022.

17. Calder PC, Ahluwalia N, Brouns F, Buetler T, Clement K, Cunningham K, et al. Dietary factors and low-grade inflammation in relation to overweight and obesity. The British journal of nutrition 2011;106 Suppl 3:S5–78 doi 10.1017/s0007114511005460.

18. Estruch R, Ros E, Salas-Salvado J, Covas MI, Corella D, Aros F, et al. Primary prevention of cardiovascular disease with a Mediterranean diet. The New England journal of medicine 2013;368(14):1279–90 doi 10.1056/NEJMoa1200303.

19. Estruch R. Anti-inflammatory effects of the Mediterranean diet: the experience of the PREDIMED study. The Proceedings of the Nutrition Society 2010;69(3):333–40 doi 10.1017/s0029665110001539.

20. Castro F, Sweeney NW, Derkach A, Traore K, Anuraj A, Guttentag L, et al. Microbial Changes in Response to a Plant-Based Diet and/or Supplements in SMM Patients: A National Multi-Arm Randomized Prospective Telehealth Study Via Healthtree: The Nutrition Prevention (NUTRIVENTION-2) Study. Blood 2022;140(Supplement 1):13079–81 doi 10.1182/blood-2022-160241.

21. Shah UA, Maclachlan KH, Derkach A, Salcedo M, Barnett K, Caple J, et al. Sustained Minimal Residual Disease Negativity in Multiple Myeloma is Associated with Stool Butyrate and Healthier Plant-Based Diets. Clinical cancer research : an official journal of the American Association for Cancer Research 2022;28(23):5149–55 doi 10.1158/1078-0432.Ccr-22-0723.

22. Shah UA, Alicea D, Adintori PA, Burge M, Blaslov J, Derkach A, et al. A Pilot Plant-Based Dietary Intervention in Overweight and Obese Patients with Monoclonal Gammopathy of Undetermined Significance and Smoldering Multiple Myeloma-the Nutrition Prevention (NUTRIVENTION) Study. Blood 2021;138:4759 doi https://doi.org/10.1182/blood-2021-151049.

23. Bottcher MR, Marincic PZ, Nahay KL, Baerlocher BE, Willis AW, Park J, et al. Nutrition knowledge and Mediterranean diet adherence in the southeast United States: Validation of a field-based survey instrument. Appetite 2017;111:166–76 doi 10.1016/j.appet.2016.12.029.

24. Krebs-Smith SM, Pannucci TE, Subar AF, Kirkpatrick SI, Lerman JL, Tooze JA, et al. Update of the Healthy Eating Index: HEI-2015. J Acad Nutr Diet 2018;118(9):1591–602 doi 10.1016/j.jand.2018.05.021.

25. Adams E, Wandro S, Avelar-Barragan J, Oliver A, Whiteson K. 2020 Low Volume Methodology for Nextera DNA Flex Library Prep Kit (96 Samples). <https://www.protocols.io/view/low-volume-methodology-for-nextera-dna-flex-librar-dm6gpr2r8vzp/v1>.

26. Beghini F, McIver LJ, Blanco-Míguez A, Dubois L, Asnicar F, Maharjan S, et al. Integrating taxonomic, functional, and strain-level profiling of diverse microbial communities with bioBakery 3. elife 2021;10:e65088 doi 10.7554/eLife.65088.

27. Oksanen J, Blanchet FG, Friendly M, Kindt R, Legendre P, McGlinn D, et al. Vegan R package: Community Ecology Package 2019.

28. Pinheiro J, Bates D, DebRoy S, Sarkar D. nlme: linear and nonlinear mixed effects models, v3.1-148 2020.

29. Avelar-Barragan J, Mendez Luque LF, Nguyen J, Nguyen H, Odegaard AO, Fleischman AG, et al. Characterizing the microbiome of patients with myeloproliferative neoplasms during a Mediterranean diet intervention. bioRxiv 2023:2023.01.25.525620 doi 10.1101/2023.01.25.525620.

30. Martinez-Gonzalez MA, Garcia-Arellano A, Toledo E, Salas-Salvado J, Buil-Cosiales P, Corella D, et al. A 14-item Mediterranean diet assessment tool and obesity indexes among high-risk subjects: the PREDIMED trial. PloS one 2012;7(8):e43134 doi 10.1371/journal.pone.0043134.

31. Emanuel RM, Dueck AC, Geyer HL, Kiladjian J-J, Slot S, Zweegman S, et al. Myeloproliferative neoplasm (MPN) symptom assessment form total symptom score: prospective international assessment of an abbreviated symptom burden scoring system among patients with MPNs. Journal of Clinical Oncology 2012;30(33):4098–103 doi 10.1200/JCO.2012.42.3863.

32. Bailey MA, Holscher HD. Microbiome-Mediated Effects of the Mediterranean Diet on Inflammation. Advances in Nutrition 2018;9(3):193–206 doi 10.1093/advances/nmy013.

33. Martín-Peláez S, Mosele JI, Pizarro N, Farràs M, de la Torre R, Subirana I, et al. Effect of virgin olive oil and thyme phenolic compounds on blood lipid profile: implications of human gut microbiota. European journal of nutrition 2017;56(1):119–31 doi 10.1007/s00394-015-1063-2.

34. Luisi MLE, Lucarini L, Biffi B, Rafanelli E, Pietramellara G, Durante M, et al. Effect of Mediterranean Diet Enriched in High Quality Extra Virgin Olive Oil on Oxidative Stress, Inflammation and Gut Microbiota in Obese and Normal Weight Adult Subjects. Front Pharmacol 2019;10:1366 doi 10.3389/fphar.2019.01366.

