## Supplemental Figures for "The NUTRIENT Trial (NUTRitional Intervention among myEloproliferative Neoplasms): Feasibility Phase"

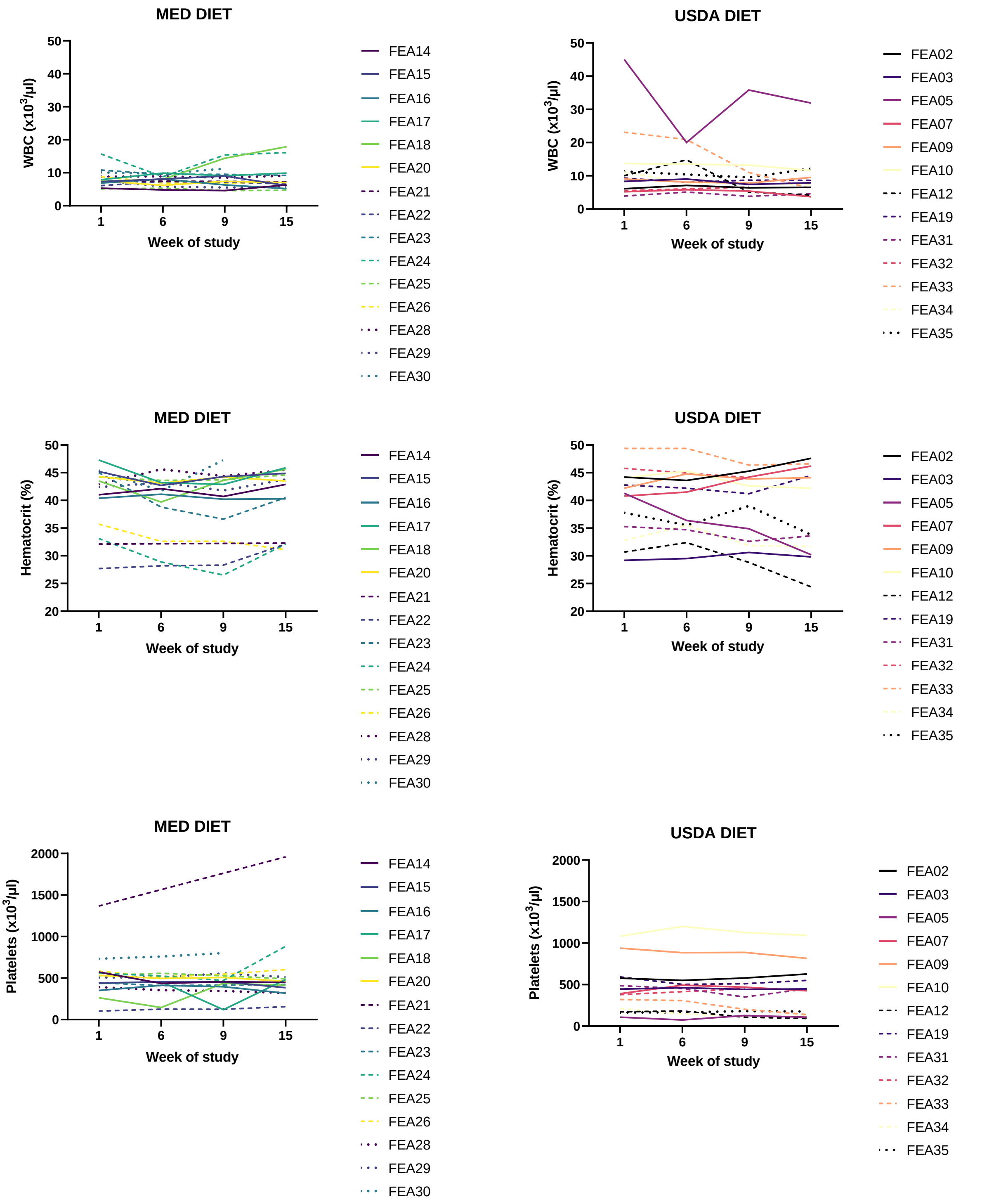


**Supplemental Figure 1.** Complete blood count (CBC) data from participants. CBC's were collected at weeks 1, 6, 9, and 15 from participants to monitor for changes in blood counts.

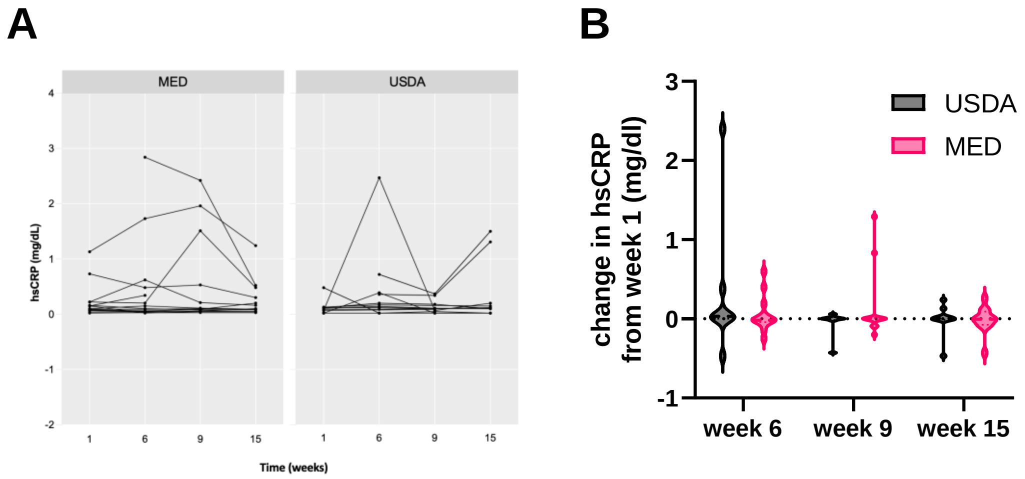


**Supplemental Figure 2. Changes in hsCRP during the study. A)** Spaghetti plot demonstrating raw hsCRP values for each participant during the course of the study, **B)** Violin plot depicting change in hsCRP at weeks 6, 9, and 15 using week 1 as baseline.

**Supplemental Figure 3. Changes in *JAK2*$^{V617F}$ allele burden over time.** Whole blood from (A) USDA and (B) MED diet groups was subjected to digital PCR to quantify *JAK2*$^{V617F}$ allele burden.

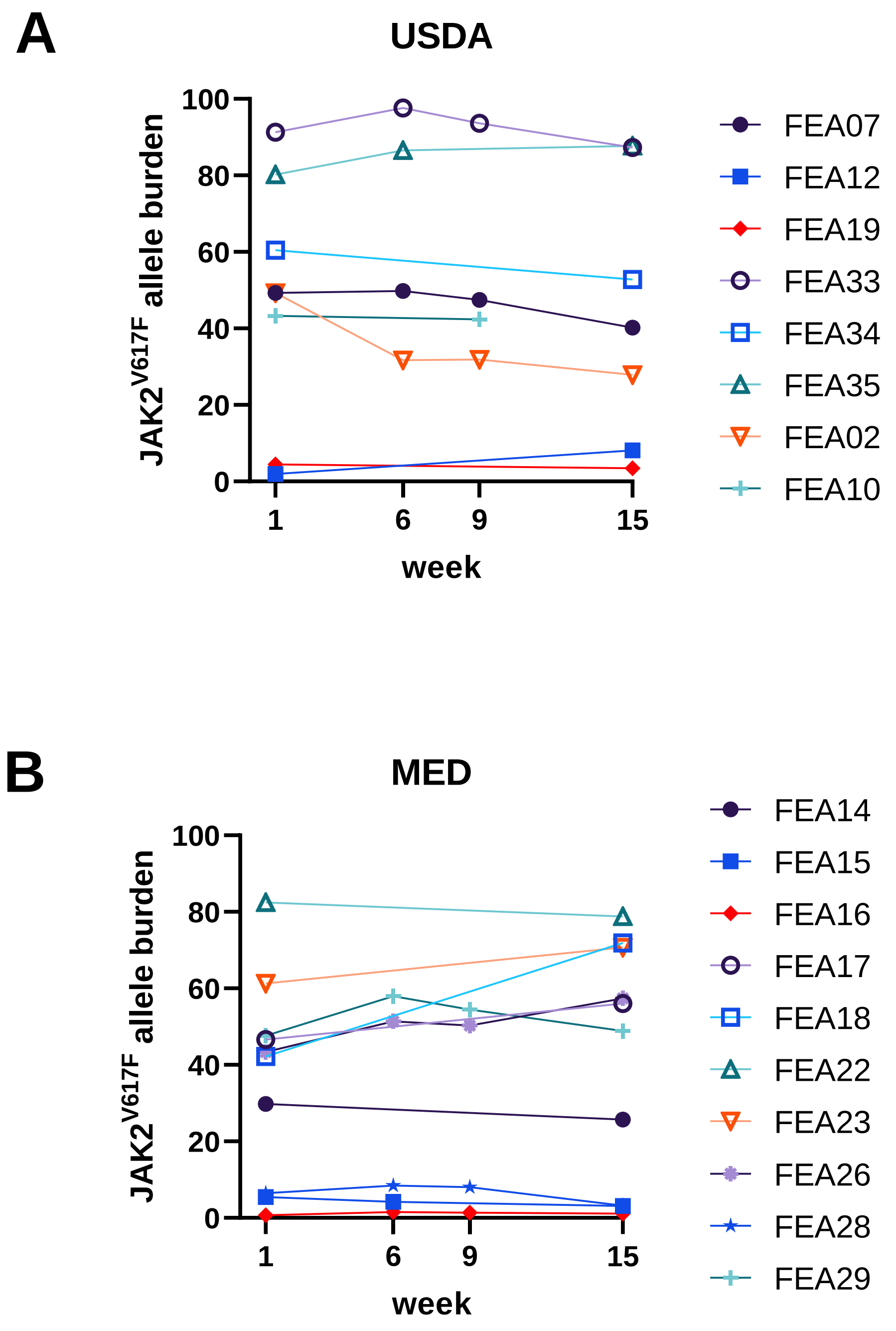

**Supplemental Table 1 – Inclusion and Exclusion Criteria**

**Inclusion Criteria**

- Age > 18 with a diagnosis of a Philadelphia negative Myeloproliferative Neoplasm including Essential Thrombocythemia (ET), Polycythemia Vera (PV), or myelofibrosis (MF)
- Any type of MPN directed therapy is allowed
- EGOG performance status of <2
- Life expectancy > 20 weeks
- Has an email address and can access the internet
- Able to read and understand English

**Exclusion Criteria**

- Pregnant or planning to become pregnant over the course of the study
- Weight loss of more than 10 pounds or 10% of the total body weight over the last 6 months
- History of allergic reactions attributed to nuts or olive oil

**Supplemental Table 2.** Longitudinal Complete Metabolic Panel Results for study cohort

| | Diet Group | Week | Na | K | Cl | CO2 | Glu | BUN | Cr | GFR | Ca | Pro | Alb | AP | AST | ALT | Bili |
|---|---|---|---|---|---|---|---|---|---|---|---|---|---|---|---|---|---|
| FEA02 | USDA | 1 | 137 | 4 | 102 | 27 | 95 | 11 | 1 | >60 | 9.5 | 7.7 | 4.7 | 48 | 21 | 14 | 1.6 |
| | | 6 | 137 | 3.9 | 104 | 24 | 102 | 16 | 1.1 | >60 | 9.5 | 7.7 | 4.6 | 54 | 23 | 18 | 1.3 |
| | | 9 | 139 | 4 | 104 | 27 | 91 | 12 | 1 | >60 | 9.3 | 7.6 | 4.5 | 56 | 22 | 16 | 1.1 |
| | | 15 | 137 | 3.6 | 103 | 27 | 93 | 15 | 1.2 | 60 | 9.5 | 7.6 | 4.8 | 55 | 30 | 24 | 1.5 |
| FEA03 | USDA | 1 | 139 | 4.2 | 104 | 26 | 97 | 17 | 0.8 | >60 | 9.6 | 6.9 | 4.4 | 52 | 19 | 11 | 0.3 |
| | | 6 | 139 | 4.5 | 103 | 27 | 97 | 20 | 0.8 | >60 | 9.4 | 7 | 4.3 | 60 | 18 | 13 | 0.3 |
| | | 9 | 139 | 4.4 | 104 | 27 | 99 | 21 | 0.7 | >60 | 9.6 | 6.8 | 4.4 | 52 | 17 | 14 | 0.3 |
| | | 15 | 139 | 4.3 | 105 | 27 | 91 | 24 | 0.7 | >60 | 9.8 | 6.8 | 4.5 | 55 | 17 | 12 | 0.3 |
| FEA05 | USDA | 1 | 137 | 4.2 | 104 | 25 | 101 | 19 | 1 | 56 | 9.9 | 6.9 | 4 | 54 | 57 | 26 | 2.6 |
| | | 6 | 135 | 4 | 103 | 25 | 106 | 12 | 0.8 | >60 | 9.1 | 7.3 | 3.3 | 48 | 41 | 19 | 2.1 |
| | | 9 | 137 | 3.8 | 104 | 27 | 101 | 10 | 0.8 | >60 | 9.3 | 7.4 | 3.4 | 51 | 51 | 23 | 2.3 |
| | | 15 | 133 | 4.2 | 102 | 25 | 88 | 8 | 0.6 | >60 | 8.9 | 6.9 | 3.4 | 50 | 47 | 24 | 2.2 |
| FEA07 | USDA | 1 | 136 | 3.8 | 102 | 26 | 94 | 13 | 0.6 | >60 | 9 | 7.5 | 3.9 | 140 | 37 | 50 | 0.6 |
| | | 6 | 138 | 4.1 | 105 | 27 | 81 | 14 | 0.7 | >60 | 9.4 | 7.4 | 4.1 | 98 | 28 | 37 | 0.5 |
| | | 9 | 135 | 4 | 101 | 29 | 70 | 19 | 0.7 | >60 | 9.8 | 7.5 | 4.2 | 100 | 36 | 54 | 0.6 |
| | | 15 | 136 | 4.1 | 102 | 31 | 74 | 13 | 0.7 | >60 | 10.1 | 8.1 | 4.5 | 106 | 30 | 33 | 0.6 |
| FEA09 | USDA | 1 | 138 | 3.9 | 104 | 28 | 123 | 14 | 0.8 | >60 | 9.5 | 7.4 | 4.4 | 75 | 28 | 29 | 0.5 |
| | | 6 | 137 | 4.2 | 100 | 30 | 146 | 20 | 0.9 | >60 | 9.8 | 7.6 | 4.6 | 79 | 36 | 30 | 0.6 |
| | | 9 | 138 | 4.2 | 104 | 26 | 145 | 24 | 0.9 | >60 | 9.9 | 7.5 | 4.5 | 72 | 30 | 24 | 0.6 |
| | | 15 | 139 | 3.9 | 102 | 29 | 139 | 16 | 0.9 | >60 | 9.7 | 7.4 | 4.6 | 71 | 25 | 17 | 0.7 |
| FEA10 | USDA | 1 | 137 | 4.3 | 102 | 30 | 86 | 14 | 1 | >60 | 9.7 | 7 | 4.3 | 54 | 17 | 12 | 1 |
| | | 6 | 140 | 4.6 | 106 | 28 | 88 | 17 | 0.9 | >60 | 9.8 | 7 | 4.7 | 64 | 18 | 14 | 0.6 |
| | | 9 | 138 | 4.8 | 104 | 27 | 72 | 17 | 0.9 | >60 | 10 | 7.2 | 4.5 | 58 | 24 | 16 | 1 |
| | | 15 | 135 | 4.1 | 102 | 28 | 64 | 14 | 0.8 | >60 | 9.5 | 6.7 | 4.6 | 46 | 23 | 21 | 0.5 |
| FEA12 | USDA | 1 | 136 | 4.6 | 105 | 25 | 95 | 23 | 1.2 | 59 | 9.2 | 7.8 | 4.6 | 31 | 19 | 17 | 0.7 |
| | | 6 | 137 | 4.5 | 104 | 22 | 104 | 29 | 1.4 | 49 | 9.6 | 8.2 | 4.4 | 59 | 23 | 34 | 0.8 |
| | | 9 | 138 | 4.7 | 107 | 23 | 110 | 19 | 1.3 | 54 | 9.1 | 7.6 | 4.2 | 35 | 21 | 17 | 0.8 |
| | | 15 | 136 | 4.7 | 107 | 23 | 97 | 27 | 1.2 | 59 | 9.4 | 7.8 | 4.4 | 41 | 22 | 19 | 0.8 |

| | | | | | | | | | | | | | | | | | |
|---|---|---|---|---|---|---|---|---|---|---|---|---|---|---|---|---|---|
| FEA19 | USDA | 1 | 138 | 4.1 | 104 | 25 | 82 | 16 | 0.8 | >60 | 9.8 | 7.3 | 4.4 | 61 | 21 | 20 | 0.4 |
| | | 6 | 140 | 4 | 106 | 28 | 96 | 9 | 0.8 | >60 | 9.4 | 7.1 | 4.3 | 62 | 20 | 18 | 0.5 |
| | | 9 | 139 | 3.6 | 107 | 27 | 87 | 15 | 0.8 | >60 | 9.4 | 7.2 | 4.4 | 62 | 22 | 19 | 0.6 |
| | | 15 | | | | | | | | | | | | | | | |
| FEA31 | USDA | 1 | 126 | 3.5 | 95 | 27 | 74 | 12 | 0.5 | >60 | 9.1 | 6.9 | 4.4 | 56 | 21 | 16 | 0.4 |
| | | 6 | 128 | 4.8 | 96 | 27 | 88 | 13 | 0.54 | 98 | 9 | 6.5 | 4.1 | 66 | 16 | 12 | 0.4 |
| | | 9 | 128 | 4.6 | 97 | 27 | 110 | 15 | 0.62 | 93 | 8.8 | 6 | 4 | 59 | 18 | 14 | 0.4 |
| | | 15 | 128 | 4.8 | 95 | 25 | 113 | 16 | 0.57 | 96 | 9.3 | 6.6 | 4.3 | 62 | 15 | 13 | 0.3 |
| FEA32 | USDA | 1 | 137 | 4 | 102 | 27 | 80 | 11 | 0.7 | >60 | 9.9 | 7.1 | 4.5 | 71 | 19 | 11 | 0.7 |
| | | 6 | | | | | | | | | | | | | | | |
| | | 9 | | | | | | | | | | | | | | | |
| | | 15 | | | | | | | | | | | | | | | |
| FEA33 | USDA | 1 | 138 | 4 | 102 | 28 | 74 | 11 | 0.7 | >60 | 9.7 | 7.9 | 4.8 | 109 | 21 | 14 | 0.7 |
| | | 6 | 137 | 4.2 | 102 | 29 | 74 | 8 | 0.6 | >60 | 9.6 | 7.6 | 4.7 | 103 | 22 | 17 | 0.7 |
| | | 9 | 138 | 4 | 103 | 28 | 99 | 10 | 0.6 | >60 | 9.3 | 7.3 | 4.4 | 89 | 19 | 16 | 0.6 |
| | | 15 | 137 | 4.3 | 102 | 30 | 86 | 12 | 0.7 | >60 | 9.3 | 7.5 | 4.4 | 77 | 21 | 15 | 0.7 |
| FEA34 | USDA | 1 | 135 | 3.7 | 103 | 25 | 73 | 12 | 0.8 | >60 | 9.2 | 6.6 | 4.3 | 58 | 29 | 15 | 1.1 |
| | | 6 | 139 | 3.9 | 106 | 26 | 99 | 13 | 0.8 | >60 | 9.1 | 6.9 | 4.3 | 75 | 57 | 78 | 0.9 |
| | | 9 | 137 | 4 | 107 | 25 | 80 | 12 | 0.8 | >60 | 8.9 | 6.3 | 4.2 | 73 | 69 | 86 | 1.1 |
| | | 15 | 136 | 4.4 | 105 | 26 | 107 | 16 | 0.9 | >60 | 8.9 | 6.5 | 4.3 | 72 | 86 | 106 | 0.9 |
| FEA35 | USDA | 1 | 140 | 4 | 102 | 33 | 63 | 12 | 0.4 | >60 | 9.9 | 7.6 | 4.7 | 141 | 27 | 23 | 0.9 |
| | | 6 | 139 | 4.5 | 101 | 31 | 103 | 11 | 0.5 | >60 | 9.9 | 7.8 | 4.7 | 147 | 31 | 28 | 0.9 |
| | | 9 | 140 | 4.2 | 103 | 30 | 93 | 11 | 0.4 | >60 | 9.4 | 7 | 4.3 | 125 | 28 | 25 | 0.7 |
| | | 15 | 136 | 4.2 | 100 | 30 | 112 | 14 | 0.4 | >60 | 9.4 | 7.1 | 4.4 | 132 | 28 | 26 | 0.8 |
| FEA14 | MED | 1 | 137 | 4.3 | 102 | 26 | 9 | 74 | 10 | 0.5 | >60 | 10 | 7.9 | 4.2 | 56 | 20 | 15 |
| | | 6 | 136 | 4 | 101 | 28 | 7 | 82 | 11 | 0.5 | >60 | 9.9 | 8 | 4.5 | 51 | 19 | 14 |
| | | 9 | 137 | 3.9 | 104 | 28 | 5 | 93 | 11 | 0.5 | >60 | 9.8 | 7.9 | 4.4 | 54 | 20 | 14 |
| | | 15 | 141 | 4 | 105 | 30 | 6 | 74 | 12 | 0.5 | >60 | 9.8 | 8.1 | 4.4 | 61 | 19 | 15 |
| FEA15 | MED | 1 | 139 | 3.9 | 103 | 28 | 8 | 87 | 15 | 0.9 | >60 | 9.9 | 7.2 | 4.3 | 91 | 19 | 13 |
| | | 6 | 137 | 3.9 | 101 | 29 | 7 | 78 | 17 | 0.9 | >60 | 9.3 | 7.1 | 4.2 | 97 | 20 | 12 |

| | | | | | | | | | | | | | | | | | |
|---|---|---|---|---|---|---|---|---|---|---|---|---|---|---|---|---|---|
| | | 9 | 137 | 3.7 | 101 | 27 | 9 | 56 | 17 | 0.9 | >60 | 9.6 | 7.2 | 4.2 | 102 | 22 | 15 |
| | | 15 | 140 | 3.7 | 102 | 28 | 10 | 69 | 21 | 0.9 | >60 | 9.4 | 7.4 | 4.2 | 94 | 20 | 12 |
| FEA16 | MED | 1 | 140 | 3.9 | 103 | 31 | 6 | 86 | 15 | 0.6 | >60 | 9.9 | 6.7 | 4 | 74 | 16 | 14 |
| | | 6 | 139 | 3.8 | 103 | 31 | 5 | 92 | 20 | 0.6 | >60 | 9.8 | 6.9 | 4.3 | 65 | 16 | 13 |
| | | 9 | 140 | 4.1 | 103 | 30 | 7 | 87 | 21 | 0.6 | >60 | 9.6 | 6.6 | 4 | 80 | 16 | 15 |
| | | 15 | 141 | 4 | 104 | 30 | 7 | 75 | 14 | 0.5 | >60 | 9.1 | 6.3 | 4 | 90 | 24 | 33 |
| FEA17 | MED | 1 | 140 | 4.1 | 105 | 28 | 7 | 81 | 14 | 1 | 57 | 9.8 | 7.2 | 4.2 | 75 | 22 | 21 |
| | | 6 | 140 | 4.3 | 102 | 29 | 9 | 93 | 19 | 0.9 | >60 | 10 | 7.3 | 4.3 | 83 | 21 | 18 |
| | | 9 | 142 | 3.9 | 103 | 30 | 9 | 61 | 14 | 1 | 57 | 9.7 | 7 | 4.3 | 84 | 22 | 19 |
| | | 15 | 139 | 4.1 | 101 | 29 | 9 | 68 | 18 | 1 | 57 | 10.1 | 7.6 | 4.5 | 96 | 27 | 27 |
| FEA18 | MED | 1 | 136 | 4.5 | 104 | 26 | 6 | 100 | 14 | 1 | >60 | 9.3 | 7.1 | 4.3 | 108 | 18 | 14 |
| | | 6 | 137 | 4.3 | 105 | 24 | 8 | 86 | 14 | 0.9 | >60 | 9.4 | 7.1 | 4 | 106 | 15 | 10 |
| | | 9 | 138 | 5 | 103 | 26 | 9 | 104 | 15 | 1 | >60 | 9 | 7.1 | 4.1 | 122 | 20 | 12 |
| | | 15 | 139 | 3.9 | 100 | 27 | 12 | 37 | 19 | 1 | >60 | 9.6 | 7.3 | 4.4 | 119 | 16 | 10 |
| FEA20 | MED | 1 | 140 | 5 | 104 | 32 | 4 | 99 | 14 | 1 | >60 | 10.1 | 7 | 4.7 | 75 | 22 | 25 |
| | | 6 | 139 | 4.9 | 104 | 31 | 4 | 90 | 15 | 1.1 | >60 | 9.5 | 6.5 | 4.2 | 61 | 24 | 21 |
| | | 9 | 140 | 4.8 | 105 | 30 | 5 | 109 | 16 | 1 | >60 | 9.8 | 6.6 | 4.4 | 60 | 24 | 23 |
| | | 15 | 134 | 4.1 | 101 | 27 | 6 | 95 | 12 | 1 | >60 | 9.6 | 6.7 | 4.3 | 71 | 25 | 25 |
| FEA21 | MED | 1 | 139 | 4.6 | 105 | 27 | 7 | 91 | 8 | 0.6 | >60 | 9.3 | 7.1 | 4.3 | 39 | 20 | 16 |
| | | 6 | | | | | | | | | | | | | | | |
| | | 9 | | | | | | | | | | | | | | | |
| | | 15 | 136 | 3.7 | 101 | 28 | 7 | 83 | 15 | 0.7 | >60 | 10.7 | 7.8 | 4.7 | 38 | 43 | 44 |
| FEA22 | MED | 1 | 141 | 4.8 | 111 | 25 | 5 | 106 | 21 | 0.7 | >60 | 9.3 | 6.4 | 4.3 | 61 | 13 | 9 |
| | | 6 | 144 | 5.4 | 109 | 25 | 10 | 86 | 17 | 0.8 | >60 | 9.6 | 6.9 | 4.6 | 65 | 14 | 9 |
| | | 9 | 143 | 5.1 | 110 | 26 | 7 | 91 | 26 | 0.8 | >60 | 9.9 | 6.9 | 4.7 | 61 | 12 | 6 |
| | | 15 | 140 | 5.3 | 110 | 26 | 4 | 97 | 22 | 0.9 | >60 | 9.9 | 6.8 | 4.6 | 60 | 15 | 10 |
| FEA23 | MED | 1 | 140 | 3.5 | 102 | 32 | 6 | 77 | 19 | 0.8 | >60 | 9.8 | 7.4 | 4.7 | 90 | 37 | 34 |
| | | 6 | 141 | 3.2 | 103 | 30 | 8 | 76 | 23 | 0.8 | >60 | 9.6 | 7.2 | 4.9 | 84 | 36 | 32 |
| | | 9 | 140 | 3.7 | 104 | 31 | 5 | 85 | 15 | 0.7 | >60 | 9.5 | 6.8 | 4.7 | 90 | 55 | 59 |
| | | 15 | 140 | 4.1 | 101 | 32 | 7 | 76 | 20 | 0.8 | >60 | 10.2 | 7.3 | 4.9 | 81 | 38 | 32 |

| | | | | | | | | | | | | | | | | |
|---|---|---|---|---|---|---|---|---|---|---|---|---|---|---|---|---|
| FEA24 | MED | 1 | 139 | 4.2 | 106 | 27 | 6 | 86 | 20 | 0.7 | >60 | 9.3 | 6.3 | 4.2 | 154 | 34 | 38 |
| | | 6 | 134 | 4.4 | 101 | 27 | 6 | 105 | 20 | 0.7 | >60 | 9.6 | 6.4 | 4.4 | 164 | 48 | 51 |
| | | 9 | 135 | 4.6 | 103 | 27 | 5 | 93 | 22 | 0.6 | >60 | 9.5 | 6.7 | 4.3 | 218 | 59 | 64 |
| | | 15 | 139 | 4.4 | 106 | 27 | 6 | 102 | 17 | 0.7 | >60 | 9.4 | 6.5 | 4.3 | 188 | 44 | 47 |
| FEA25 | MED | 1 | 141 | 4.8 | 103 | 30 | 8 | 88 | 18 | 0.8 | >60 | 10.4 | 7.4 | 4.6 | 52 | 17 | 14 |
| | | 6 | 139 | 4.6 | 101 | 30 | 8 | 69 | 16 | 0.7 | >60 | 10 | 7.3 | 4.6 | 51 | 19 | 13 |
| | | 9 | 137 | 4.2 | 100 | 30 | 7 | 43 | 18 | 0.7 | >60 | 9.6 | 7.1 | 4.3 | 49 | 16 | 12 |
| | | 15 | 137 | 4.6 | 101 | 31 | 5 | 64 | 17 | 0.8 | >60 | 9.8 | 7.3 | 4.5 | 52 | 19 | 16 |
| FEA26 | MED | 1 | 141 | 4.4 | 107 | 26 | 8 | 67 | 24 | 1 | >60 | 8.9 | 6.7 | 4.3 | 65 | 23 | 23 |
| | | 6 | 139 | 4.4 | 105 | 31 | 3 | 88 | 21 | 1 | >60 | 9.4 | 6.4 | 4.4 | 53 | 17 | 15 |
| | | 9 | 141 | 0.5 | 106 | 28 | 7 | 82 | 20 | 1 | >60 | 9.2 | 6.6 | 4.4 | 56 | 20 | 14 |
| | | 15 | 138 | 4.4 | 106 | 27 | 5 | 96 | 20 | 1.1 | >60 | 9.2 | 6.3 | 4.4 | 56 | 18 | 14 |
| FEA28 | MED | 1 | 136 | 4.3 | 101 | 28 | 7 | 83 | 21 | 0.8 | >60 | 9.5 | 8.4 | 4.5 | 78 | 39 | 43 |
| | | 6 | 138 | 4.4 | 101 | 28 | 9 | 68 | 16 | 0.8 | >60 | 10.1 | 8.4 | 4.8 | 71 | 29 | 23 |
| | | 9 | 135 | 4.6 | 101 | 28 | 6 | 81 | 22 | 0.8 | >60 | 10 | 8.2 | 4.6 | 66 | 27 | 29 |
| | | 15 | 134 | 4.2 | 99 | 27 | 8 | 81 | 18 | 0.8 | >60 | 9.9 | 8.6 | 4.7 | 68 | 41 | 41 |
| FEA29 | MED | 1 | 139 | 4.2 | 103 | 28 | 8 | 80 | 21 | 1 | >60 | 9.5 | 7.3 | 4.7 | 45 | 20 | 21 |
| | | 6 | 138 | 3.9 | 103 | 29 | 6 | 67 | 19 | 0.9 | >60 | 9.7 | 6.9 | 4.5 | 46 | 26 | 32 |
| | | 9 | 139 | 4.3 | 104 | 29 | 6 | 70 | 20 | 1 | 55 | 9.5 | 7.1 | 4.8 | 41 | 22 | 23 |
| | | 15 | 142 | 4.3 | 104 | 28 | 10 | 69 | 17 | 1 | 55 | 9.8 | 7.4 | 4.7 | 39 | 26 | 25 |
| FEA30 | MED | 1 | 137 | 4 | 100 | 26 | 11 | 80 | 14 | 0.9 | >60 | 10.2 | 8.3 | 5.1 | 83 | 30 | 36 |
| | | 6 | 137 | 4.1 | 101 | 28 | 8 | 82 | 13 | 0.9 | >60 | 10.6 | 7.8 | 4.8 | 74 | 33 | 38 |
| | | 9 | | | | | | | | | | | | | | | |
| | | 15 | | | | | | | | | | | | | | | |

**Supplemental Table 3.** Longitudinal Lipid Panels for Study Cohort

| | Week | Diet Group | Tot Chol | TG | HDL | LDL | VLDL | Non-HDL |
|---|---|---|---|---|---|---|---|---|
| FEA02 | 1 | USDA | 227 | 159 | 53 | 142 | 32 | 174 |
| | 6 | | 227 | 120 | 57 | 146 | 24 | 170 |
| | 9 | | 227 | 156 | 51 | 145 | 31 | 176 |
| | 15 | | 258 | 122 | 57 | 177 | 24 | 201 |
| FEA03 | 1 | USDA | 186 | 200 | 41 | 105 | 40 | 145 |
| | 6 | | 222 | 159 | 47 | 143 | 32 | 175 |
| | 9 | | 208 | 169 | 48 | 126 | 34 | 160 |
| | 15 | | 209 | 126 | 49 | 135 | 25 | 160 |
| FEA05 | 1 | USDA | 168 | 109 | 47 | 108 | 10 | |
| | 6 | | 123 | 131 | 21 | 76 | 26 | 102 |
| | 9 | | 113 | 120 | 20 | 69 | 23 | 93 |
| | 15 | | 119 | 126 | 18 | 76 | 25 | 101 |
| FEA07 | 1 | USDA | 164 | 135 | 42 | 95 | 27 | 122 |
| | 6 | | 161 | 94 | 48 | 94 | 19 | 113 |
| | 9 | | 176 | 95 | 55 | 102 | 19 | 121 |
| | 15 | | 177 | 104 | 49 | 107 | 21 | 128 |
| FEA09 | 1 | USDA | | | | | | |
| | 6 | | 161 | 84 | 57 | 87 | 17 | 104 |
| | 9 | | 172 | 120 | 48 | 100 | 24 | 124 |
| | 15 | | 135 | 97 | 44 | 72 | 19 | 91 |
| FEA10 | 1 | USDA | 92 | 58 | 38 | 42 | 12 | 54 |
| | 6 | | 88 | 51 | 45 | 33 | 10 | 43 |
| | 9 | | 97 | 36 | 47 | 43 | 7 | 50 |
| | 15 | | 84 | 54 | 44 | 29 | 11 | 40 |
| FEA12 | 1 | USDA | | | | | | |
| | 6 | | 76 | 60 | 23 | 41 | 12 | 53 |
| | 9 | | 83 | 78 | 35 | 32 | 16 | 48 |
| | 15 | | 69 | 62 | 27 | 30 | 12 | 42 |
| FEA19 | 1 | USDA | 314 | 183 | 62 | 215 | 37 | 252 |
| | 6 | | 245 | 155 | 48 | 166 | 31 | 197 |
| | 9 | | 272 | 112 | 56 | 194 | 22 | 216 |
| | 15 | | | | | | | |
| FEA31 | 1 | USDA | 185 | 59 | 58 | 115 | 12 | 127 |
| | 6 | | | | | | | |
| | 9 | | | | | | | |
| | 15 | | | | | | | |
| FEA32 | 1 | USDA | 281 | 112 | 69 | 190 | 22 | 212 |
| | 6 | | | | | | | |
| | 9 | | | | | | | |

| | | | | | | | | |
|---|---|---|---|---|---|---|---|---|
| | 15 | | | | | | | |
| FEA33 | 1 | USDA | 218 | 353 | 59 | 88 | 71 | 159 |
| | 6 | | 200 | 125 | 56 | 119 | 25 | 144 |
| | 9 | | 204 | 220 | 58 | 102 | 44 | 146 |
| | 15 | | 196 | 169 | 66 | 96 | 34 | 130 |
| FEA34 | 1 | USDA | 117 | 157 | 51 | 35 | 31 | 66 |
| | 6 | | 105 | 81 | 44 | 45 | 16 | 61 |
| | 9 | | 110 | 88 | 50 | 42 | 18 | 60 |
| | 15 | | 105 | 89 | 48 | 39 | 18 | 57 |
| FEA35 | 1 | USDA | 138 | 381 | 26 | 36 | 76 | 112 |
| | 6 | | 145 | 160 | 33 | 80 | 32 | 112 |
| | 9 | | | | | | | |
| | 15 | | 130 | 167 | 31 | 66 | 33 | 99 |
| FEA14 | 1 | MED | 158 | 85 | 48 | 93 | 17 | 110 |
| | 6 | | 188 | 92 | 57 | 113 | 18 | 131 |
| | 9 | | 158 | 139 | 50 | 80 | 28 | 108 |
| | 15 | | 163 | 125 | 53 | 85 | 25 | 110 |
| FEA15 | 1 | MED | 233 | 80 | 64 | 153 | 16 | 169 |
| | 6 | | 196 | 87 | 52 | 127 | 17 | 144 |
| | 9 | | 214 | 174 | 57 | 122 | 35 | 157 |
| | 15 | | 240 | 83 | 61 | 162 | 17 | 179 |
| FEA16 | 1 | MED | 172 | 84 | 55 | 100 | 17 | 117 |
| | 6 | | 173 | 130 | 48 | 99 | 26 | 25 |
| | 9 | | 150 | 148 | 42 | 78 | 30 | 108 |
| | 15 | | 139 | 191 | 39 | 62 | 38 | 100 |
| FEA17 | 1 | MED | 216 | 82 | 44 | 156 | 16 | 172 |
| | 6 | | 235 | 139 | 45 | 162 | 28 | 190 |
| | 9 | | 200 | 116 | 46 | 131 | 23 | 154 |
| | 15 | | 224 | 120 | 46 | 154 | 24 | 178 |
| FEA18 | 1 | MED | 96 | 88 | 33 | 45 | 18 | 63 |
| | 6 | | 102 | 96 | 31 | 52 | 19 | 71 |
| | 9 | | 105 | 156 | 24 | 50 | 31 | 81 |
| | 15 | | 123 | 175 | 32 | 56 | 35 | 91 |
| FEA20 | 1 | MED | 168 | 150 | 39 | 99 | 30 | 129 |
| | 6 | | 150 | 178 | 40 | 74 | 36 | 110 |
| | 9 | | 153 | 172 | 41 | 78 | 34 | 112 |
| | 15 | | 149 | 126 | 39 | 85 | 25 | 110 |
| FEA21 | 1 | MED | 120 | 25 | 68 | 47 | 5 | 52 |
| | 6 | | | | | | | |
| | 9 | | | | | | | |
| | 15 | | 141 | 23 | 75 | 61 | 5 | 66 |
| FEA22 | 1 | MED | 125 | 141 | 27 | 70 | 28 | 98 |

| | | | | | | | | |
|---|---|---|---|---|---|---|---|---|
| | 6 | | 125 | 111 | 28 | 75 | 22 | 97 |
| | 9 | | 124 | 80 | 26 | 82 | 16 | 98 |
| | 15 | | 130 | 111 | 28 | 80 | 22 | 102 |
| FEA23 | 1 | MED | 251 | 74 | 94 | 142 | 15 | 157 |
| | 6 | | 214 | 79 | 84 | 114 | 16 | 130 |
| | 9 | | 206 | 71 | 83 | 109 | 14 | 123 |
| | 15 | | 223 | 96 | 97 | 107 | 19 | 126 |
| FEA24 | 1 | MED | 217 | 107 | 77 | 119 | 21 | 40 |
| | 6 | | 207 | 98 | 66 | 121 | 20 | 141 |
| | 9 | | 209 | 71 | 65 | 130 | 14 | 144 |
| | 15 | | 205 | 110 | 60 | 123 | 22 | 145 |
| FEA25 | 1 | MED | 211 | 141 | 69 | 114 | 28 | 142 |
| | 6 | | 199 | 120 | 66 | 109 | 24 | 133 |
| | 9 | | 202 | 101 | 63 | 119 | 20 | 139 |
| | 15 | | 203 | 97 | 62 | 122 | 19 | 141 |
| FEA26 | 1 | MED | 142 | 78 | 43 | 83 | 16 | 99 |
| | 6 | | 139 | 132 | 37 | 76 | 26 | 102 |
| | 9 | | 148 | 129 | 42 | 80 | 26 | 106 |
| | 15 | | 129 | 73 | 43 | 71 | 15 | 86 |
| FEA28 | 1 | MED | 204 | 104 | 49 | 134 | 21 | 155 |
| | 6 | | 194 | 96 | 47 | 128 | 19 | 147 |
| | 9 | | 201 | 205 | 45 | 115 | 41 | 156 |
| | 15 | | 204 | 127 | 50 | 129 | 25 | 154 |
| FEA29 | 1 | MED | 178 | 207 | 43 | 94 | 41 | 135 |
| | 6 | | 163 | 233 | 42 | 74 | 47 | 121 |
| | 9 | | 189 | 200 | 42 | 107 | 40 | 147 |
| | 15 | | 170 | 242 | 41 | 81 | 48 | 129 |
| FEA30 | 1 | MED | 150 | 154 | 49 | 70 | 31 | 101 |
| | 6 | | 137 | 177 | 54 | 48 | 35 | 83 |
| | 9 | | | | | | | |
| | 15 | | | | | | | |
